# Real-time spatial health surveillance: mapping the UK COVID-19 epidemic

**DOI:** 10.1101/2020.08.17.20175117

**Authors:** Richard Fry, Joe Hollinghurst, Helen R Stagg, Daniel A Thompson, Claudio Fronterre, Chris Orton, Ronan A Lyons, David V Ford, Aziz Sheikh, Peter J Diggle

**Affiliations:** Health Data Research, UK; Medical School, Lancaster University; Swansea University Medical School; Usher Institute, University of Edinburgh

## Abstract

The COVID-19 pandemic has highlighted the need for robust data linkage systems and methods for identifying outbreaks of disease in near real-time. Using self-reported app data and the Secure Anonymised Information Linkage (SAIL) Databank, we demonstrate the use of sophisticated spatial modelling for near-real-time prediction of COVID-19 prevalence at small-area resolution to inform strategic government policy areas. A pre-requisite to an effective control strategy is that predictions need to be accompanied by estimates of their precision, to guard against over-reaction to potentially spurious features of ‘best guess’ predictions. In the UK, important emerging risk-factors such as social deprivation or ethnicity vary over small distances, hence risk needs to be modelled at fine spatial resolution to avoid aggregation bias. We demonstrate that existing geospatial statistical methods originally developed for global health applications are well-suited to this task and can be used in an anonymised databank environment, thus preserving the privacy of the individuals who contribute their data.

## 1 Introduction

On 11th March 2020, the World Health Organization declared a pandemic of COVID-19 caused by the SARS-CoV-2 coronavirus [1]. By this date, the UK had reported 373 confirmed COVID-19 cases and six deaths [2] Up to 15^th^ July 2020, these figures had risen to 291,911 and 45,053 [3]. Lockdowns governing the movement of the population and opening of shops and other facilities, initially imposed across the entire country on 23^rd^ March 2020 [4], have been a key tool in the government’s response to COVID-19. Since that date, detection of local variations in infection rates has been critical for controlling the spread of SARS-CoV-2 [5], including ascertaining the level of required local public health response across the UK. A key example of this was the implementation of the first ‘local lockdown’ in Leicester on 30th June 2020, in response to a cluster of COVID-19 accounting for approximately one in ten of all new disease cases across the country in the preceding week [6]

The COVID Symptom Study app (Zoe Global Limited, King’s College London) was released publicly on 24^th^ March 2020[7], the day after the UK-wide lockdown rules were first imposed. The app collects postcode of residence at the time of registration and daily updates on self-reported COVID-19-associated symptoms. The Secure Anonymised Information Linkage (SAIL) Databank facilitates robust secure storage and use of anonymised person-based data for research to improve health, well-being and services [8, 9]. During the pandemic, SAIL has been receiving daily updates of the COVID Symptom Study app data, facilitating near real-time health surveillance of COVID-19 across the UK.

To help understand the localised spread and flare up of the disease we adapted existing statistical methodology [10] for the analysis of geo-referenced health outcome data to map, at Lower-layer Super Output Area (LSOA) resolution (Datazone in Scotland and Super Output Area in Northern Ireland), the prevalence of positive symptom reports amongst app users over a rolling 14-day period, together with associated limits of statistical uncertainty. Notwithstanding the limitations of this self-reported health outcome, these maps provide the first fine-scale, UK-wide assessment of the geographical distribution of probable COVID-19 infections, and have been used by the devolved administrations in each country for pandemic planning [11][12].

## 2 Methods

### 2.1 Population Sampling

Across the UK, as of 15^th^ July 2020, 4,002,397 individuals had registered on the COVID Symptom Study app. Use of the app is voluntary and thus the population sample is non-random. Users of the app had to have access to an internet-enabled telephone, although reporting for multiple individuals in the same household was instigated on 1^st^ May 2020 for those people who could not access or use the app [13]. At the time of registration, individuals report baseline demographic and clinical information (e.g. underlying health conditions), as well as postcode of residence. Self-reported data on COVID-19-associated symptoms, including fever and persistent cough, are recorded for any day on which an individual reports. For a full metadata summary see the HDR Gateway deposit [14].

The SAIL Databank acts as a secure gateway to the ZOE app data for the whole of the UK. Data are made available daily via a secure data transfer and processed into a SQL DB2 database. Access to the data is via a secure remote desktop login following approval for a project via an application to the SAIL Information Governance Review Panel (IGRP). In light of the COVID-19 crisis, IGRP applications were typically approved within 24-hours. Prior to transfer of the data to the SAIL Databank, postcode data were aggregated to LSOA level using the Office for National Statistics (ONS) postcode lookup directory [15] to maintain an app user’s privacy.

### 2.2 Informatics

Data were extracted from the SAIL Databank using SQL and processed to generate suitable inputs for the geospatial modelling. Where an individual reported their symptoms more than once in a day, the last record was taken. Likely instances of COVID-19 were calculated through either a) the presence of high fever and persistent cough (’classic symptoms’) or b) an algorithm developed by the King’s College team, which used an array of symptoms and other characteristics (persistent cough, skipping meals, loss of smell, gender, age and fatigue; ‘multi-symptom algorithm’), [16]. The denominator of users in each LSOA for each analysis was calculated using a 14-day retrospective window i.e. the number of individuals who had reported data to the app at any time during that period. The numbers of app users and cases were then aggregated to the LSOA level. The resulting data for each LSOA consisted of its population-weighted centroid, *x*, the number of people who used the app at least once over the time-period in question, *n*, and the number of those who were predicted to have COVID-19 at least once within the time-period, *y*.

### 2.3 Geospatial statistical model and inference

Our statistical model is a geospatial extension of the logistic regression model for binomial (numerator/denominator) data, in which the log-odds of the probability, *P*(*x*), of at least one positive symptom report is the unobserved realisation of a spatially correlated stochastic process and, conditional on *P*(*x*), the corresponding numerator *y* follows a binomial distribution with denominator *n*. The model has three parameters that determine the mean and variance of *P*(*x*) and the rate at which the correlation between the values of *P*(*x*) at two different locations decays with increasing distance between them.

We estimated the parameters using Monte Carlo maximum likelihood and used the fitted model to draw samples from the joint predictive distribution of *P*(*x*) over all LSOA population weighted centroids. Parameters were re-estimated separately for each of the UK’s constituent countries, in each case using data aggregated over a rolling 14-day time-period. In the Supplementary Material we describe in detail how we developed and fitted the particular model that we used for our application to the COVID-19 app data.

The complete prediction for each LSOA is a probability distribution for its underlying prevalence. This distribution can be summarised as the user wishes. We chose to map four summaries: the mean, a point prediction of prevalence; the 5% and 95% quantiles, which together measure the uncertainty associated with each point prediction; and the probability that the prevalence in the LSOA in question is greater than the country-wide average for each devolved nation, with mapped values close to 1 or 0 indicating “hot-spots” and “cold-spots” respectively. If the primary aim of the mapping is hot-spot detection, additional useful summaries would be the probabilities that local prevalence exceeds each of a set of thresholds representing increasing multiples of the country-wide average. Patches of mapped probabilities close to 1 would then indicate both the geographical extent and magnitude of local hot-spots. Predicted prevalence data were summarised for the whole of the UK, and for each of its four constituent countries.

## 3 Results

### 3.1 App users

Table 1 summarises the users of the app for the UK as of 14^th^ July 2020. Across the UK users were predominately female, white, between 30 and 55 and lived in the least deprived areas. It is also worth noting that the 20% of users did not provide a full postcode thereby limiting the utility of these users data provision for the purposes of high-resolution spatial modelling.

**Table 1:** Summary of registered ZOE Symptom Study App users as of 14^th^July 2020.

|  | Number | Percentage |
| --- | --- | --- |
| **Registered App Users** | 3840824 | 100% |
| **Age** |  |  |
| Median (IQR) | 40 (30,55) |  |
| 0-10 | 154091 | 4.0% |
| 15-20 | 346591 | 9.0% |
| 25-30 | 692117 | 18.0% |
| 35-40 | 714001 | 18.6% |
| 45-50 | 682677 | 17.8% |
| 55-60 | 536308 | 14.0% |
| 65-70 | 321197 | 8.4% |
| 75-80 | 85531 | 2.2% |
| 85+ | 21204 | 0.6% |
| Not Recorded | 307360 | 8.0% |
| **Gender** |  |  |
| Female | 2150324 | 56.0% |
| Male | 1380425 | 35.9% |
| Prefer not to say | 2458 | 0.1% |
| intersex | 256 | 0.1% |
| Not Recorded | 307361 | 8.0% |
| **Ethnicity** |  |  |
| Hispanic | 52 | 0.1% |
| Other | 11883 | 0.3% |
| Prefer not to say | 10322 | 0.3% |
| Asian (UK) | 56140 | 1.5% |
| Black (UK) | 15935 | 0.4% |
| Chinese | 9134 | 0.2% |
| Middle Eastern | 10154 | 0.3% |
| Mixed (other) | 30974 | 0.8% |
| Mixed (White/Black) | 16051 | 0.4% |
| White (UK) | 2448755 | 63.8% |
| US Residents | 1151 | 0.1% |
| Not Recorded | 1230273 | 32.0% |
| **Deprivation** |  |  |
| Townsend Quintile (where full postcode provided at registration) | 3067197 | 79.9% |
| 1. Least deprived | 814557 | 26.6% |
| 2 | 741201 | 24.2% |
| 3 | 629180 | 20.5% |
| 4 | 490686 | 16.0% |
| 5. Most deprived | 391573 | 12.8% |

### 3.2 App usage over time

There was no national government requirement for members of the public to use the app. In each country, the number of people registering to use the system increased rapidly in the early weeks of its availability, and more slowly thereafter (Figure 1, top panel). In England, Scotland and Wales the number of active users (people who recorded one or more app submission in the preceding 14 days) also increased between mid-April and early May, but declined thereafter, from a peak of around 60% in early May to around 45% in mid July. (Figure 1, bottom panel). In Northern Ireland, where the app is, in effect, competing with Northern Ireland’s own app [17] the percentage of active users peaked at around 50% in early May and had declined to about 30% by mid July.

**Figure 1:**
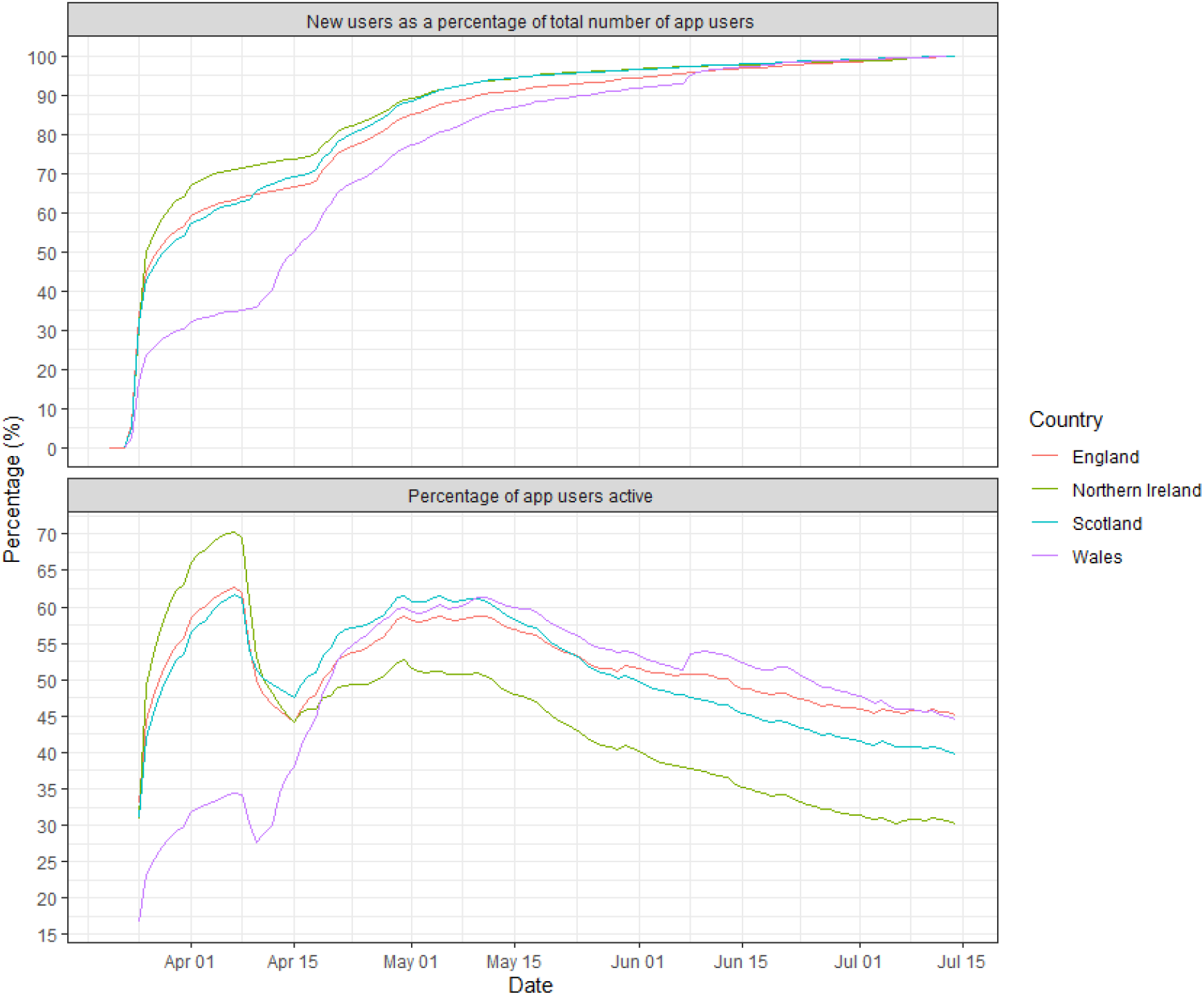
Longitudinal trajectories of app use for each of the four UK countries up to 14^th^July 2020, as percentage of total registered app users by 14^th^ July 2020 (upper panel) and as percentage active within a fourteen-day rolling time-window (lower panel)

### 3.3 Predicted prevalence of COVID-19 over time

Predicted disease prevalence over time, weighted for population size within each LSOA, was plotted using both classic symptoms and the multi-symptom algorithm. Both provided similar patterns of predicted disease prevalence for the first two weeks of data collection (Figure 2), the figures from the multi-symptom algorithm were higher than those using classic symptoms (classic symptoms 3.6% to 4.3% across the four countries at their peak, multi-symptom algorithm 6.0% to 7.5%) but diverged thereafter. Using classic symptoms, predicted prevalence slowly declined to mid-May and then was approximately stable at slightly more than 2% in Wales, slightly less than 2% in England, Scotland and Northern Ireland (Figure 2, upper panel). Using the multi-symptom algorithm, the decline was more rapid, stabilising around two weeks earlier at approximately 0.4% in all four countries. (Figure 2, lower panel)

**Figure 2:**
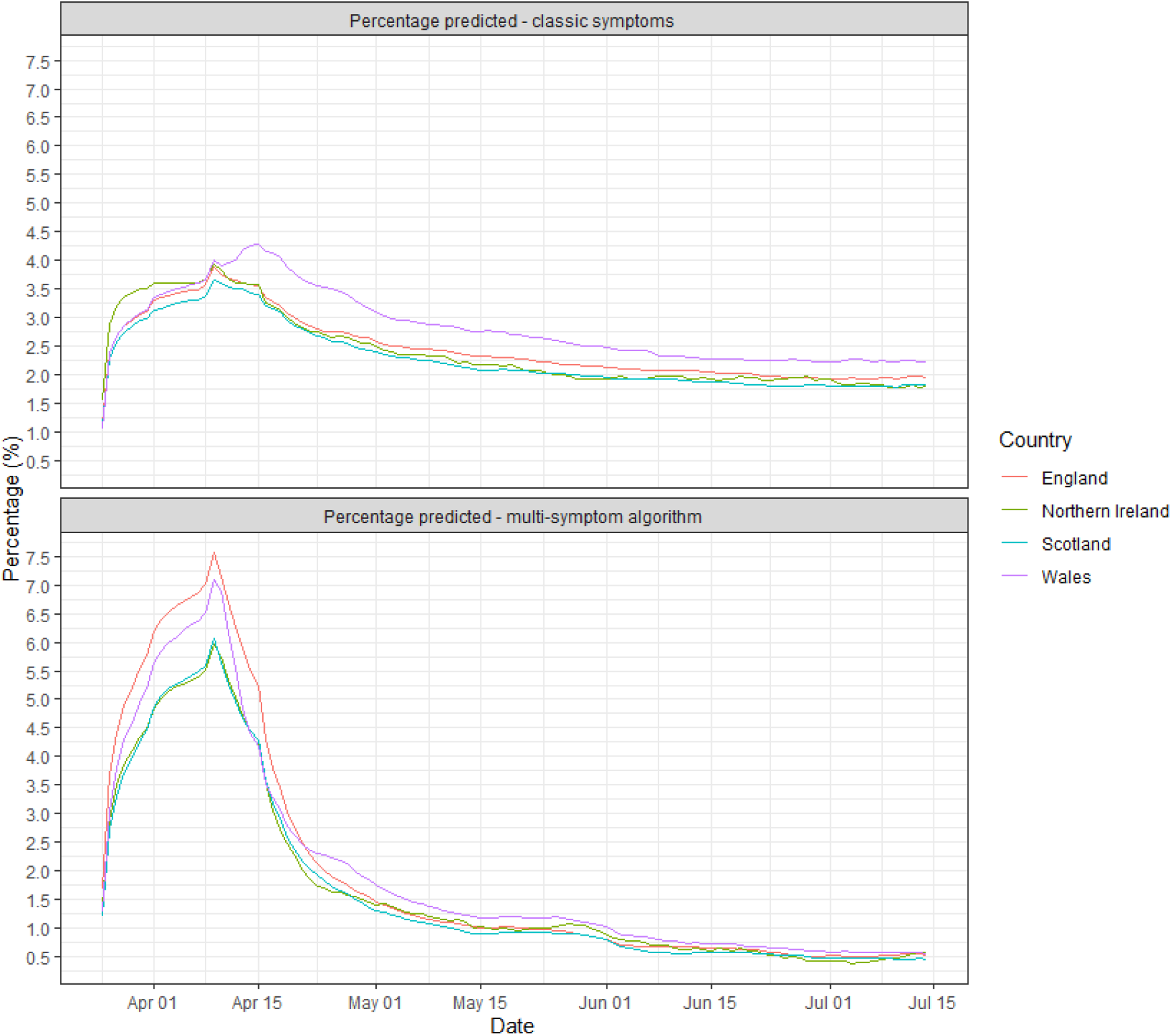
Longitudinal trajectories of symptom-prevalence (percentage of active users reporting symptoms) for each of the four UK countries up to 14^th^ July 2020, based on classic symptoms (upper panel) and multi-symptom algorithm (lower panel).

### 3.4 Predictive mapping

Predictive mapping at LSOA-level geography based on inputs derived using the prevalence algorithm described in Menni et al.[16] revealed small-scale spatial variation in disease prevalence, which varied over time. Figures 3 and 4 show UK-wide LSOA-level maps of predicted prevalence and predictive probability that each LSOA exceeded the national average prevalence, over the pandemic to 15^*th*^ July 2020. Most hot-spots (bright yellow areas in Figure 4) were located in or close to major cities, with Aberdeen and Bristol as notable exceptions.

**Figure 3:**
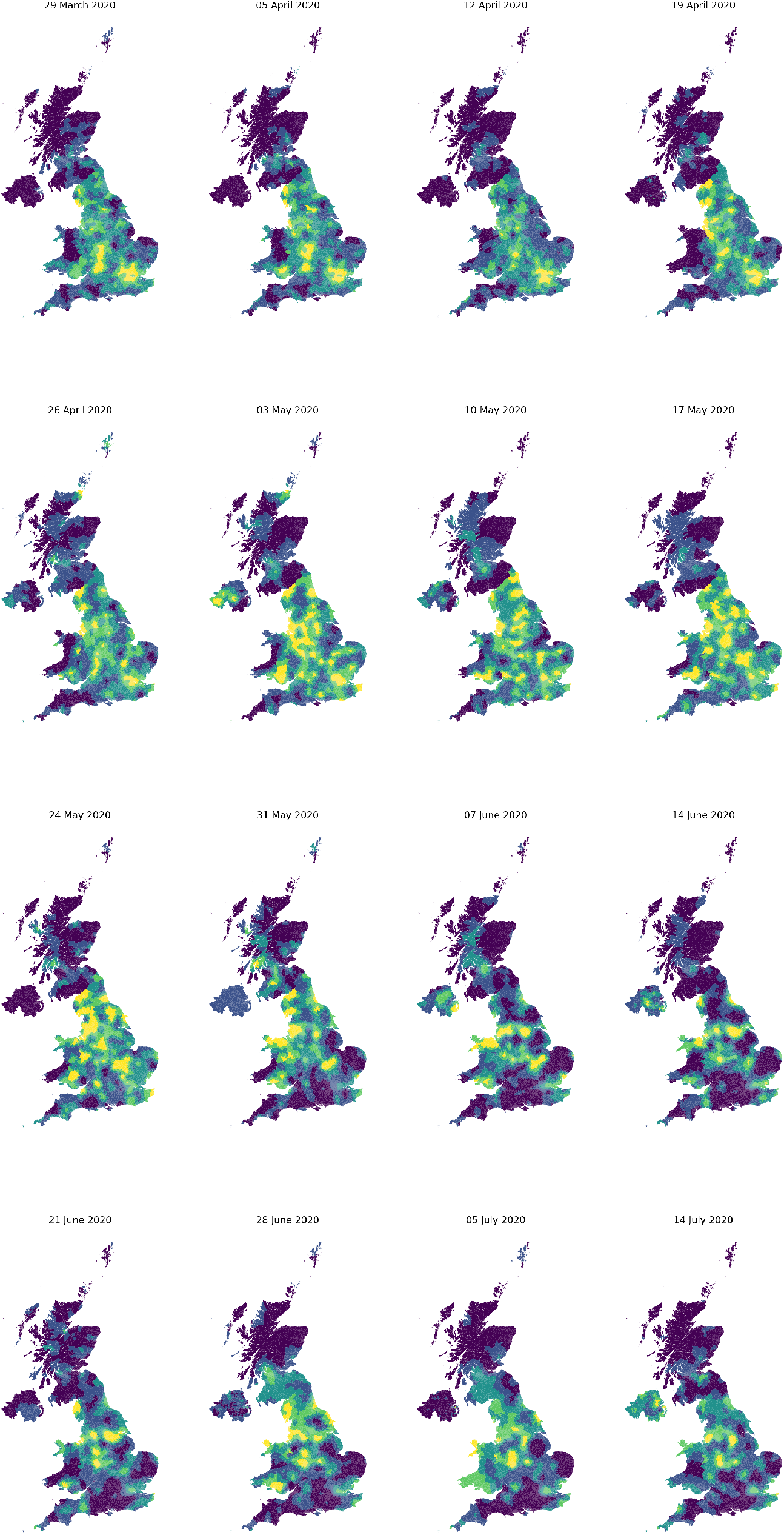
LSOA Level COVID-19 prevalence predictions using a 14-day window 9^*th*^ March - 14^*th*^ July 2020. Purples represent low values, green mid range and yellow high values.

**Figure 4:**
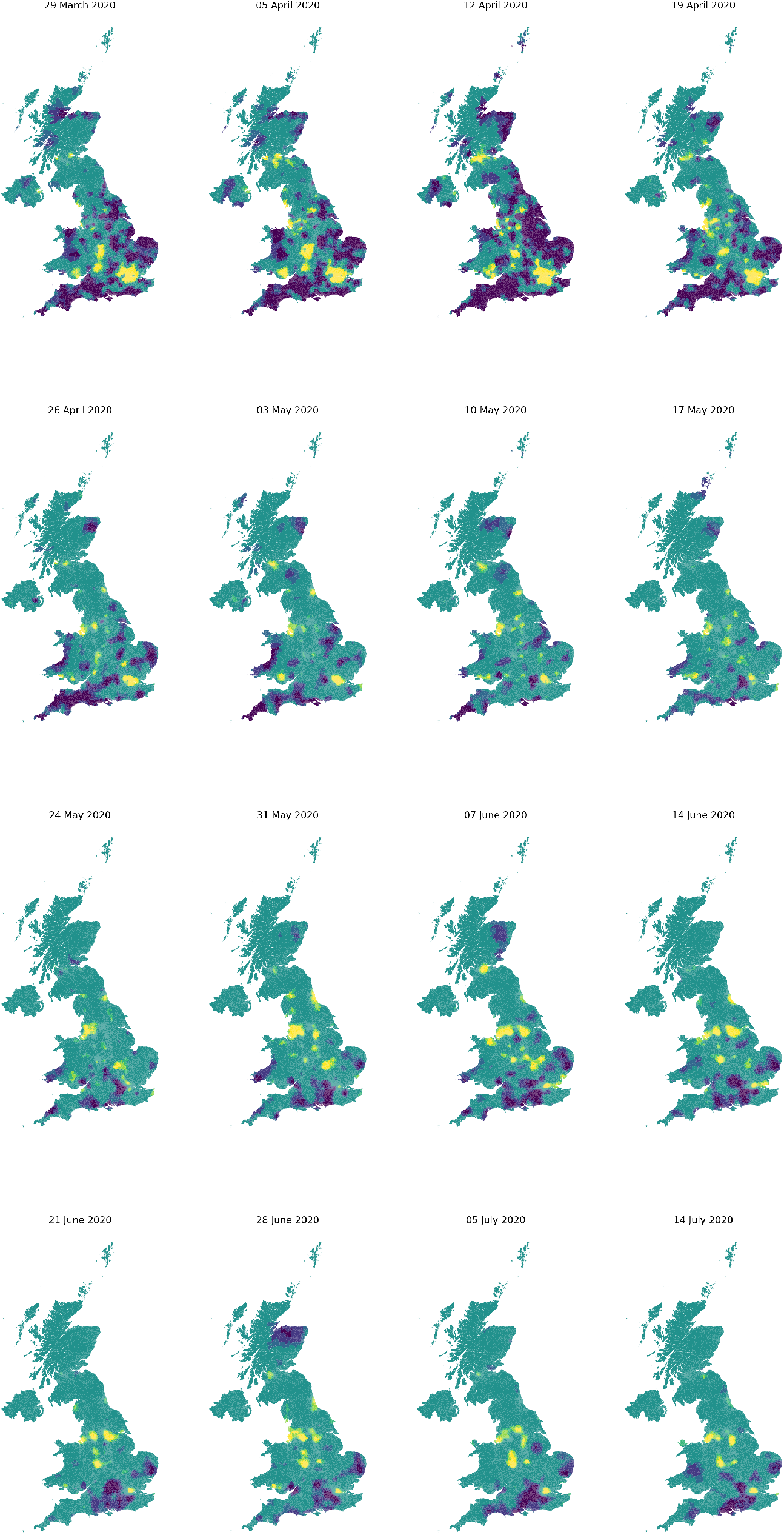
Predictive probabilities for LSOA-level prevalence to exceed the UK-wide average using a 14-day window 29^*th*^ March - 14^*th*^ July 2020. Purples represent low probabilities, green mid range and yellow high probabilities.

Prevalence and exceedance probability maps need to be interpreted in combination. The prevalence exceedance (hot-spot) maps focus attention on areas that show statistically significant deviations above the national averages. However, all other things being equal, these were likely to occur in areas that have high population density and, consequently, deliver more precise local predictions. The three-panel format of Figure 5 faciliates this combined interpretation by allowing the reader to check whether areas indicated in Figure 4 as hot-spots are also areas whose predicted prevalence is markedly high, and conversely, whilst the left-hand and right-hand panels act as a guard against over-interpretation of imprecise point predictions. For example, the centre-panel of Figure 5 shows that the largely rural area of west Cumbria had relatively high prevalence over the 14-day period ending 28^*th*^ June, but the associated probability limits were wide, and the bottom row, second-left column of Figure 4 does not indicate any part of west Cumbria to have been a hot-spot. Conversely, over this same period, prevalence levels in London were no longer among the highest in the country (Figure 5, centre panel), but were nevertheless almost certainly above the English national average (Figure 4, bottom row, second-left column).

**Figure 5:**
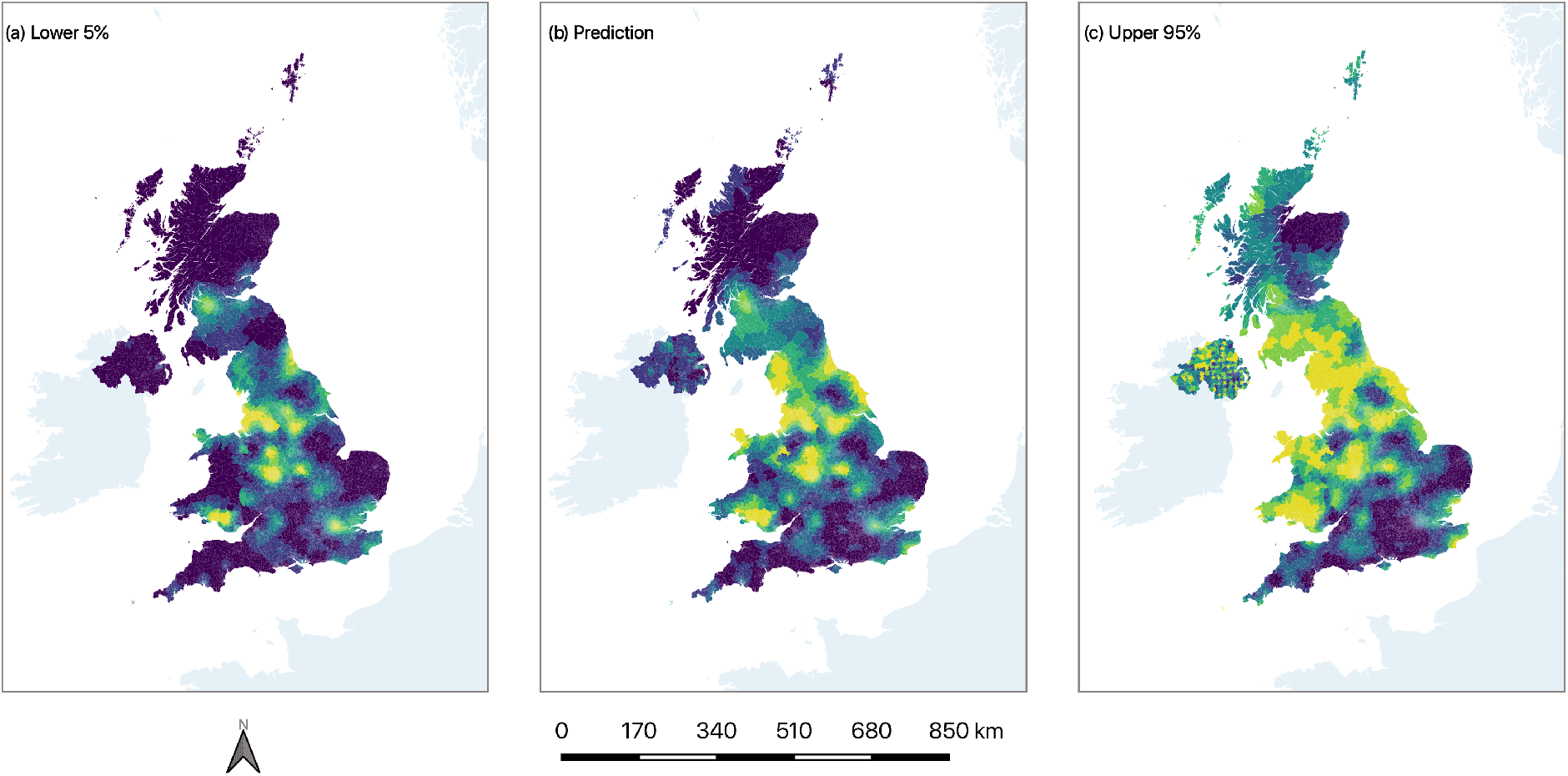
Predicted (centre panel (b)), lower 5% (left-hand panel (a)) and upper 95% (right-hand panel (c)) limits for the probability that LSOA-level prevalence to exceed the UK-wide average over the 14-day period ending 28^*th*^ June 2020. Purples represent low values, green midrange and yellow high values.

## 4 Discussion

The COVID-19 pandemic has shown how a combination of skills (Health Informatics, Statistics and Geography) can provide insights to inform local and national government policy at a UK level. Building on the HDR UK ‘One Institute’ principles, we have prototyped and delivered data infrastructures and analysis pipelines capable of delivering timely and insightful analytics to all levels of government. These have formed a cornerstone of the COVID-19 response in Scotland and Wales in particular, and illustrate how fine-grained spatio-temporal inferential mapping tools for near-real-time crowd-sourced georeferenced health outcome data are critical to inform rapid public health responses.

The platform established within the SAIL system should not be seen as uniquely applicable to data from the COVID Symptom Study app, or indeed to COVID-19. Such tools can be applied to any source of georeferenced health data, including when comparing data sourced via different detection and testing platforms, each of which may have a different target population, sensitivity and specificity. We note the particular relevance to newly emerging infectious conditions – where spatiotemporal mapping of spread is critical – and thus the need to maintain such platforms so that they can be activated as each pandemic arises, e.g. for influenza[18]. The required geospatial methodology for these tools has been available since the early years of the century [19]. Diggle, Rowlingson and Su [20] report on a real-time surveillance system for calls to the now-defunct NHS Direct on-line triage service [21] for which the primary reason for the call was recorded as non-specific gastro-intestinal illness. This system was developed in collaboration with the Southampton Public Health Service and ran in prototype form over the years 2001 to 2003.

There has been a proliferation of mapped outputs related to the COVID-19 pandemic with policy makers and the general public equally concerned with tracking the pandemic. However, most of the published maps have been at a regional level [7, 5, 22]. Although valuable, these can mask localised hotspots as exemplified by the Leicester, UK outbreak. In situations where expedient decisions are required, mapping statistical outputs to granular geographies, with supporting information relating to confidence intervals, gives policy makers better information to take necessary action. Also, by recognising and exploiting spatial correlation in the underlying prevalence surface, geospatial statistical methods can deliver substantially more precise estimation of local prevalence than classical methods that implicitly assume independence of outcomes in different spatial units [23]

We acknowledge the limitations that come with using self-reported symptom data from an app used voluntarily. Firstly, confirmation that self-reported symptoms did indeed represent COVID-19 disease was not possible at a UK level, although the multi-symptom algorithm utilised was generated using predictive regression modelling comparing symptoms to self-reported reverse transcription polymerase chain reaction SARS-CoV-2 test results [16]. Secondly, the individuals included in the studied population are not a random sample of the UK population, potentially presenting a source of collider bias due to the link between age and app usage[24], nor are they necessarily representative with respect to other factors that are either known, or thought likely, to affect susceptibility; for example, gender or ethnicity. The inclusion in the model of LSOA-level covariate information is a potential route to controlling for these at LSOA level, although not at individual level. For example, in the current COVID-19 context information on the age distribution of app users would allow adjustment for the potentially non-representative sub-population of active app users. For environmentally driven health outcomes, such as asthma symptom exacerbation in relation to air quality, covariate adjustment could also materially improve predictive precision.

This combination of real-time data sources and rapid analytical tools using readily adaptable methodologies is a powerful one in the control of pathogens where evolving spatio-temporal patterns of incidence are of public health concern. Our response to COVID-19 has much to teach us about preparedness for the next pandemic[18, 25]

## 5 Conclusions

In conclusion, we have demonstrated the value of a real-time spatio-temporal inferential mapping platform for public health efforts during the emergence and spread of infectious diseases. The work has been conducted in the confines of the privacy-protecting SAIL Databank to produce statistically robust results at a spatially granular level whilst ensuring that no individual contributor to the ZOE Symptom Study app can be identified. Such tools are not only essential to produce population-weighed estimates of disease prevalence, but they provide a unique insight into the geographical distribution of the disease, thus informing local and national control efforts.

## Data Availability

All data made available via the Secure Anonymised Information Linkage (SAIL) Databank

https://saildatabank.com

## 6 Acknowledgements

This work uses data provided by participants of the COVID-19 Symptoms Study, developed by ZOE Global Limited with scientific and clinical input from King’s College London. We would also like to acknowledge all data providers who made anonymised data available for research.

We wish to acknowledge the collaborative partnership that enabled acquisition and access to the de-identified data, which led to this output. The collaboration was led by BREATHE – The Health Data Research Hub for Respiratory Health, in partnership with SAIL Databank at Swansea University, the Health Data Research UK Swansea University site team and the Usher Institute at the University of Edinburgh. We wish to acknowledge the input of ZOE Global Limited and King’s College London in their development and sharing of the data, and their input into the understanding and contextualisation of data for COVID-19 research. All research conducted has been completed under the permission and approval of SAIL independent Information Governance Review Panel (IGRP) project number 1078. HRS is supported by the Medical Research Council [MR/R008345/1].

## Supplementary material: Geospatial statistical model and inference

### S.1 Model formulation

The standard way to relate prevalence – the probability of a positive outcome – to one or more variable risk-factors is a logistic regression equation. For example, writing *P*(*x*) for the probability of a positive outcome at a location *x* within a specified time-interval, if geographical variation in prevalence were thought to be primarily associated with deprivation, a candidate logistic regression model would be log[*P*(*x*)*/{*1 *− P*(*x*)*}*] = *α* + *β ×* deprivation score

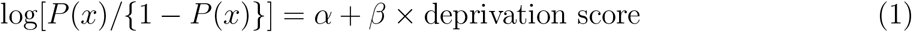

In a geospatial model, equation (1) is extended to

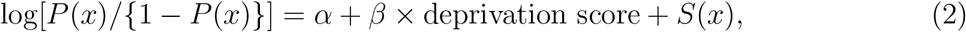

where now *S*(*x*) is an unobserved, spatially correlated stochastic process. Its role is to account for that part of the geographical variation in prevalence that is *not* explained by available covariates. Its inclusion in the model has two potentially important consequences: it guards against invalid inferences on covariate effects by acknowledging that data from different locations may not be statistically independent; more importantly in the current context, it delivers more precise estimates of the underlying prevalence surface *P*(*x*) by transferring some of what would otherwise have been treated as unpredictable random variation into predictable, spatially correlated variation. The extent to which these potential benefits are realised in practice is context-specific – the stronger the spatial correlation structure, the greater the benefits. This makes it essential that the spatial correlation structure is estimated from the data, rather than being pre-specified.

### S.2 Exploratory analysis

In preliminary analyses of data from different parts of the UK we did not find a robust association of app-reported symptom prevalence with LSOA-level deprivation or population density. We therefore fitted a version of the geospatial model (2) without covariate-adjustment, i.e. all geographical variation in app-reported symptom prevalence is ascribed to the stochastic term *S*(*x*).

To explore the spatial correlation structure of the data, we converted the positive symptom report count, *y*, and the number of active users, *n*, for each LSOA in a given 14-day time-window into empirical logits, *el* = log{(*y* + 0.5)*/*(*n − y* + 0.5)*}* and calculated their *variogram*, as follows. Associate each empirical logit, *el_i_*, with the population-weighted centroid, *x_i_*, of the corresponding LSOA. Calculate the quantities 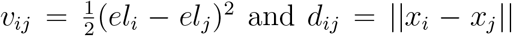, the distance between *x_i_* and *x_j_*. Group the *d_ij_* in a set of distance bins centred on *u_k_*: *k* = 1*,…, m* and calculate 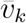 as the average of all of the *v_ij_* whose corresponding *d_ij_* fall within the *k*th distance bin. A plot of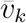 against *u_k_*, called the *empirical variogram*, estimates the quantity *V* (*u_k_*) = *τ*^2^+*σ*^2^{1*−ρ*(*u*), where *σ*^2^ is the variance of *S*(*x*), *ρ*(*u*) is the correlation between pairs *S*(*x*) and *S*(*x*′) at locations a distance *u* apart, and *τ*^2^ > 0 represents the sampling variation in the individual *el_i_*. Figure 6 shows the result using square-root-transformed empirical logits from Scotland’s data over the 14-day time-window 1 to 14 April 2020.

**Figure 6:**
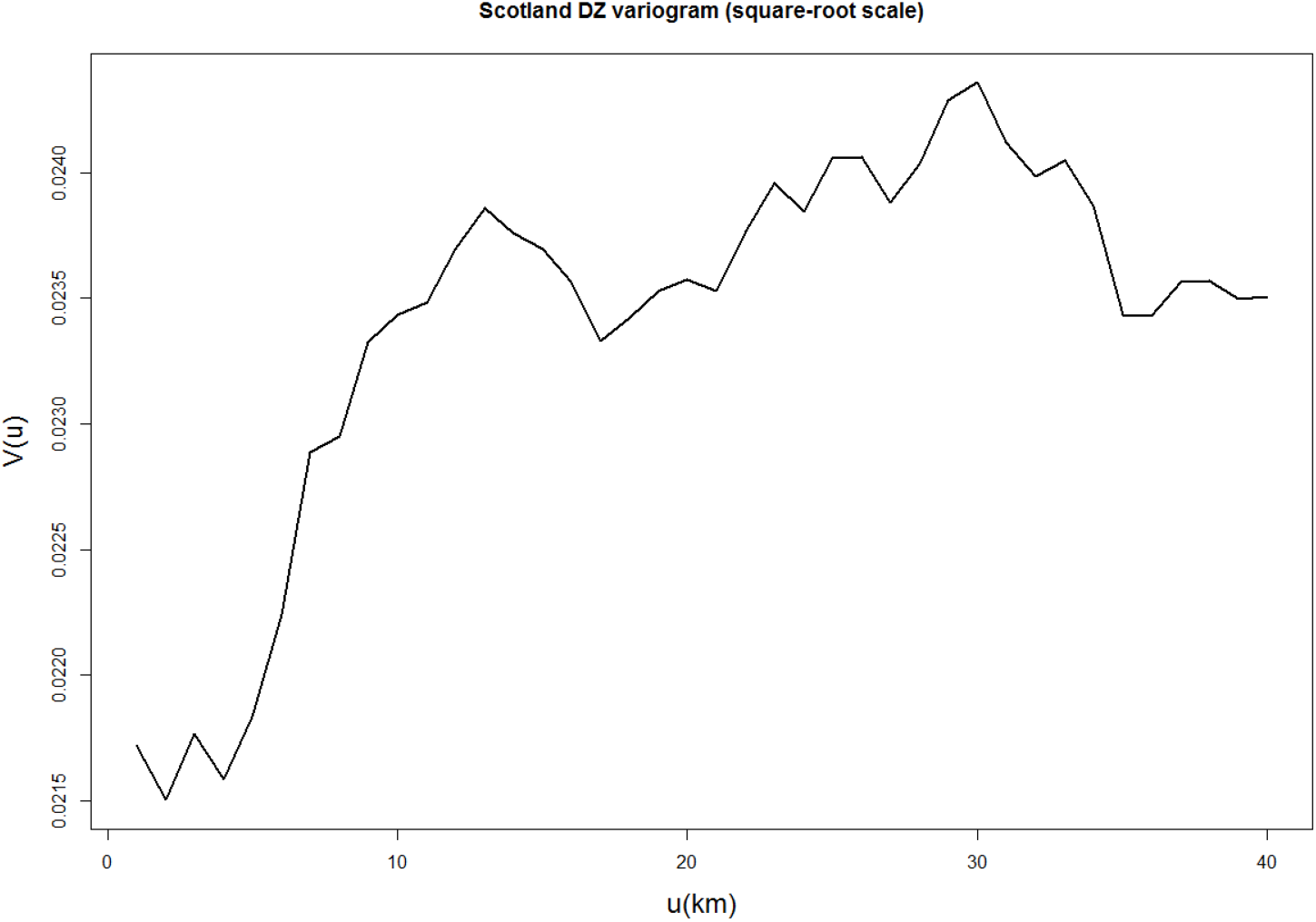
Empirical variogram for empirical logits of positive symptom reports in Scotland over the 14-day time-window 1 to 14 April 2020.

Based on Figure 6 and similar results from other parts of the UK we used a standard specification for *S*(*x*) as a Gaussian process with twice-differentiable Matérn correlation function (Diggle and Giorgi, 2019, p32), giving a model with three parameters: the intercept, *α*, in (2), the variance, *σ*^2^, of the stochastic process *S*(*x*), which represents how variable are the log-odds of prevalence from place to place; and a correlation parameter, *φ*, which determines the rate at which the correlation between log-odds of prevalence at two locations decays as the distance between them increases.

### S.3 Inference

We estimated the model parameters by Monte Carlo maximum likelihood, as implemented in the `R` package `PrevMap` (Giorgi and Diggle, 2017). This method ensures statistical efficiency, but requires the tuning of a Markov chain Monte Carlo algorithm. We ran the chain with 11,000 samples, from which the first 1,000 were discarded and the remainder sampled every tenth iteration, leaving a sample of size 1,000 from the joint predictive distribution of the prevalences *P*(*x*) for each LSOA. Initial values of the model parameters were taken from visual inspection of the empirical variogram shown in Figure 6, but optimised separately for each time-window of data in each of the UK’s constituent countries.

## Summary Points

What is known:

- COVID-19 has highlighted the need for robust methods for identifying outbreaks of disease and local levels
- Most mapping efforts have so far been restricted to regional level estimates - there are very few local level estimates of COVID-19 prevalence
- Self reported app data is currently being contributed by 4 million people in the UK

What we are adding:

- We demonstrate the use of sophisticated spatial modelling for near-real-time prediction of COVID-19 prevalence at small-area resolution to inform strategic government policy areas
- We provide estimates of their precision, to guard against over-reaction to potentially spurious features of ‘best guess’ predictions
- We demonstrate that adapting existing geospatial statistical methods, originally developed for global health applications, can be used in an anonymised databank environment, thus preserving the privacy of the individuals who contribute their data.

## References

[1] World Health Organization. WHO Timeline - COVID-19. URL: https://www.who.int/news-room/detail/08-04-2020-who-timeline---covid-19.

[2] World Health Organization. COVID-19 situation reports. URL: https://www.who.int/emergencies/diseases/novel-coronavirus-2019/situation-reports.

[3] Coronavirus (COVID-19) in the UK. URL: https://coronavirus.data.gov.uk/.

[4] Prime Minister’s statement on coronavirus (COVID-19): 23 March 2020 - GOV.UK. URL: https://www.gov.uk/government/speeches/pm-address-to-the-nation-on-coronavirus-23-march-2020.

[5] Hagai Rossman et al. A framework for identifying regional outbreak and spread of COVID-19 from one-minute population-wide surveys. May 2020. doi: 10.1038/s41591-020-0857-9. URL: https://doi.org/10.1038/s41591-020-0845-0.

[6] Plans for managing the coronavirus (COVID-19) outbreak in Leicester – GOV.UK. URL: https://www.gov.uk/government/speeches/local-action-to-tackle-coronavirus.

[7] COVID Symptom Study – Help slow the spread of COVID-19. URL: https://covid.joinzoe.com/.

[8] Ronan A. Lyons et al. “The SAIL databank: Linking multiple health and social care datasets”. In: BMC Medical Informatics and Decision Making 9.1 (Jan. 2009), pp. 1–8. issn: 14726947. doi: 10.1186/1472-6947-9-3. URL: https://link.springer.com/articles/10.1186/1472-6947-9-3%20 https://link.springer.com/article/10.1186/1472-6947-9-3.

[9] David V. Ford et al. “The SAIL Databank: Building a national architecture for e-health research and evaluation”. In: BMC Health Services Research 9 (2009), p. 157. issn: 14726963. doi: 10.1186/1472-6963-9-157.

[10] Peter Diggle and Emanuele Giorgi. Model-based geostatistics for global public health: methods and applications. isbn: 9781138732353.

[11] Welsh Government. New app launched to track and trace coronavirus. 2020. url: https://media.service.gov.wales/news/new-app-launched-to-track-and-trace-coronavirus.

[12] Technical Advisory Cell: summary of advice 5 June 2020 — GOV.WALES. URL: https://gov.wales/technical-advisory-cell-summary-advice-5-june-2020.

[13] Charities back COVID Symptom Tracker. URL: https://covid.joinzoe.com/post/charity-release.

[14] Health Data Research UK. ZOE Metadata. URL: https://metadata-catalogue.org/hdruk/#/catalogue/dataModel/06f8c66d-4e91-44dc-a109-1df729b72b61/properties.

[15] Office for National Statistics. ONS Postcode Directory (May 2019) — Open Geography portal. URL: http://geoportal.statistics.gov.uk/datasets/ons-postcode-directory-may-2019.

[16] Cristina Menni et al. “Real-time tracking of self-reported symptoms to predict potential COVID-19”. In: Nature Medicine (May 2020), pp. 1–4. issn: 1546170X. doi: 10.1038/s41591-020-0916-2.

[17] Coronavirus (COVID-19): overview and advice — nidirect. URL: https://www.nidirect.gov.uk/articles/coronavirus-covid-19-overview-and-advice#toc-2.

[18] Colin R. Simpson et al. The UK hibernated pandemic influenza research portfolio: triggered for COVID-19. July 2020. doi: 10.1016/S1473-3099(20)30398-4. URL: https://doi.org/10.1016/S1473-3099.

[19] Anders Brix and Peter J. Diggle. “Spatiotemporal prediction for log-Gaussian Cox processes”. In: Journal of the Royal Statistical Society: Series B (Statistical Methodology) 63.4 (Nov. 2001), pp. 823–841. issn: 1369-7412. doi: 10.1111/1467-9868.00315. url: http://doi.wiley.com/10.1111/1467-9868.00315.

[20] Peter Diggle, Barry Rowlingson, and Ting-li Su. “Point process methodology for on-line spatio-temporal disease surveillance”. In: Environmetrics 16.5 (Aug. 2005), pp. 423–434. issn: 1180-4009. doi: 10.1002/env.712. URL: http://doi.wiley.com/10.1002/env.712.

[21] National Symptom Surveillance Using Calls to a Telephone Health Advice Service — United Kingdom, December 2001–February 2003. URL: https://www.cdc.gov/mmwr/preview/mmwrhtml/su5301a33.htm.

[22] The Path to Zero: Key Metrics For COVID Suppression – Pandemics Explained. URL: https://globalepidemics.org/key-metrics-for-covid-suppression/.

[23] Claudio Fronterre et al. “Design and Analysis of Elimination Surveys for Neglected Tropical Diseases”. In: The Journal of Infectious Diseases 221.Supplement 5 (Jan. 2020), S554–S560. issn: 0022-1899. doi: 10.1093/infdis/jiz554. URL: https://doi.org/10.1093/infdis/jiz554.

[24] Gareth Griffith et al. “Collider bias undermines our understanding of COVID-19 disease risk and severity”. In: medRxiv (May 2020), p. 2020.05.04.20090506. doi: 10.1101/2020.05.04.20090506. URL: https://www.medrxiv.org/content/10.1101/2020.05.04.20090506v2.

[25] Honglei Sun et al. “Prevalent Eurasian avian-like H1N1 swine influenza virus with 2009 pandemic viral genes facilitating human infection”. In: Proceedings of the National Academy of Sciences (June 2020), p. 201921186. issn: 0027-8424. doi: 10.1073/pnas.1921186117. URL: http://www.pnas.org/lookup/doi/10.1073/pnas.1921186117.

## References

Diggle, P.J. and Giorgi, E. (2019). Model-based Geostatistics: Methods and Applications in Global Public Health. Boca Raton: CRC Press

Giorgi, E. and Diggle, P.J. (2017). PrevMap: an R package for prevalence mapping. Journal of Statistical Software, 78, 1–29, doi:10.18637/jss.v078.i08

